# Cytokines IL-1β, TNF-α, and IL-10 as Possible Biomarker Molecules Predictive of Sepsis

**DOI:** 10.1101/2025.04.19.25326102

**Authors:** Linda M. Chams, Rafael Guillermo Villarreal Julio, Luis David German, María F. Yasnot, Gleymer Morelo Espitia, Manuel Camilo Meneses, Gabriel Pérez, Carlos J. Castro

## Abstract

**Introduction:** sepsis causes a high percentage of morbidity and mortality in patients treated in the ICU, it is a complex pathology both in its definition and in its pathophysiology, with major anachronisms in the diagnosis, based on signs, symptoms and laboratory data that do not provide sufficient information to direct treatment, for this reason it is necessary to implement a diagnostic method to determine the patient’s septic and prognostic status.

**Objective:** to determine the expression of the cytokines IL-1, TNF and IL-10 in patients admitted to the Intensive Care Unit (ICU) of a Hospital in Montería, Córdoba.

**Materials and methods:** descriptive study, with 40 ICU patients who met the inclusion criteria. The serum levels of the cytokines were measured with Elisa and the results were analyzed with GraphPad Prism 6, applying the U Mann Whitney test for non-parametric data.

**Results:** the plasma IL-10 concentration was of great relevance in septic patients, being elevated in the pulmonary focus, the main generator of sepsis.

**Conclusion:** the cytokine IL-10 was considered a possible predictor of mortality in patients with sepsis.

## INTRODUCTION

Sepsis is associated with a high mortality rate (70%) in hospitalized patients, with percentages close to those associated with acute myocardial infarction (1). According to the WHO and PAHO, approximately 31 million people experience an episode of sepsis each year, and of these, approximately 6 million people die (1).

In Latin America, higher mortality rates have been recorded than in European countries and the United States, and it is noted with concern that the most affected Latino population is those under 50 years of age. This behavior may possibly be related to risk factors due to the living conditions to which people are exposed in these developing countries (1). Sepsis induces the immune system to react to eliminate the pathogen that is causing it, for this reason, its severity is not only determined by the viability of the invading pathogen that can be toxic and tissue-destructive, but also by the host response, which can be exaggerated and cause collateral damage to tissues and organs, since highly potent effectors do not discriminate between pathogenic targets and their own (2-4). Sepsis continues to be defined by nonspecific physiological alterations caused by cellular processes that lead to acute or exacerbated organ dysfunction (5-8).

Therefore, an uncertain diagnosis leads to a delay in the initiation of antimicrobial therapy, which can influence mortality in certain age groups of patients with severe sepsis. Therefore, the use of alternative diagnostic tools such as biomarkers is essential. Biomarkers are biological parameters that provide information about the normal or pathological state of an individual or a population, and are used to understand different diseases in various aspects such as: treatment, prevention, diagnosis and progression of the disease and responses to therapy, in addition to evaluation of therapeutic intervention (9-11)

The use of biomarkers as an early diagnosis would allow for rapid treatment, improving the chances of recovery of patients and helping to reduce unnecessary antibiotic therapy (6) (7). The search for specific sepsis biomarkers may become a good diagnostic alternative to determine the host response and the detection of pathogens, helping to optimize the management of patients with sepsis (9) (10).

Based on the above, the objective of this research is to evaluate the cytokines IL-1β, TNFα, and IL-10 as potential predictive biomarker molecules for sepsis in patients in the Intensive Care Unit (ICU) of a hospital in the city of Montería. Likewise, it is intended to open the door to future research, where other molecules are evaluated as potential biomarkers for sepsis in the study population, and at the same time, achieve a complete characterization that allows reflecting the conditions and variations of these.

## MATERIALS AND METHODS

A descriptive study was conducted with patients admitted to the adult ICU at a hospital in the city of Montería-Córdoba, who met the inclusion criteria of the study, between September 2015 and March 2016.

Forty ICU patients were selected without discrimination based on sex or age. Of these, 20 were diagnosed with sepsis and the other 20 were patients with pathologies other than sepsis. The clinical sample used was patient plasma, obtained by centrifuging EDTA-anticoagulated whole blood, separating it into vials within 5 hours of collection and storing them at -20°C.

The inclusion criteria were as follows: patients whose relatives signed the informed consent where they were informed of the objective of the study, patients with a medical diagnosis of sepsis or septic shock who presented some or several of the following parameters: temperature: >38°C or <36°C, Leukocytosis:

>12,000 μL or Leukopenia: <4,000 μL, normal white blood cell count with more than 10% of immature forms, blood pressure: Systolic blood pressure

<90 mmHg or Mean arterial pressure <70 mmHg, heart rate: >90/min and respiratory rate:

>20/min.

Patients selected for the study who met the inclusion criteria were provided with medical records, which included patient identification, clinical information, signs and symptoms, laboratory data, and treatment. Invasive procedures, which could lead to septic status, were highlighted as important data. Patients who were on mechanical ventilation were distinguished from those who were not.

For the determination of serum levels of TNF-α, IL-1β and IL-10, the commercial kit LEGEND MAX™ Human TNF-α ELISA Kit, Human IL-1β and Human IL-10 Biolegend (San Diego, CA) was used respectively, according to established procedures.

by the kit. In summary, the method consisted of a quantitative determination by solid-phase capture ELISA, where the antibodies specific for human TNF-α, IL-1β and IL-10 are attached.

The antibody allows the binding and immobilization of the respective antigens (TNF-α, IL-1β and IL-10). Subsequently, a second antibody was bound to a different site of the antigen, after removing the excess antibody the Avidin-HRP B, Avidin-HRP D, Avidin-HRP A solution was added, for TNF-α, IL-1β and IL-10 respectively, which bound to the antibody added previously. After this binding, the excess was washed and the substrate was added with which a colored compound was obtained, after the appropriate time, stopping solution was added and finally it was read at 450 nm.

Regarding the results analysis, a descriptive study was conducted with the collected samples, which were grouped into a case group, those diagnosed with sepsis, and a control group, those hospitalized patients who did not present a diagnosis of sepsis. Both groups underwent statistical analysis, obtaining means, standard deviations, and percentages. The data obtained were classified as nonparametric using the Shapiro-Wilk test, and the groups were compared using the Mann-Whitney U test. In addition, a calibration curve was created to determine the concentrations of the cytokines IL-1, TNF, and IL-10.

## RESULTS

The mean age in the sepsis group was 56.45 +/- 24.52 years, while in the non-sepsis group it was 48.15 +/- 23.47 years (p=0.3578). Of the 40 participants, 55% were women and the remaining 45% were men, of whom 50% had sepsis. In the sepsis group, 60% were women and 40% were men; while in the non-sepsis group, there was an equal distribution of men and women (50%). See Table 1.

**Table 1.**
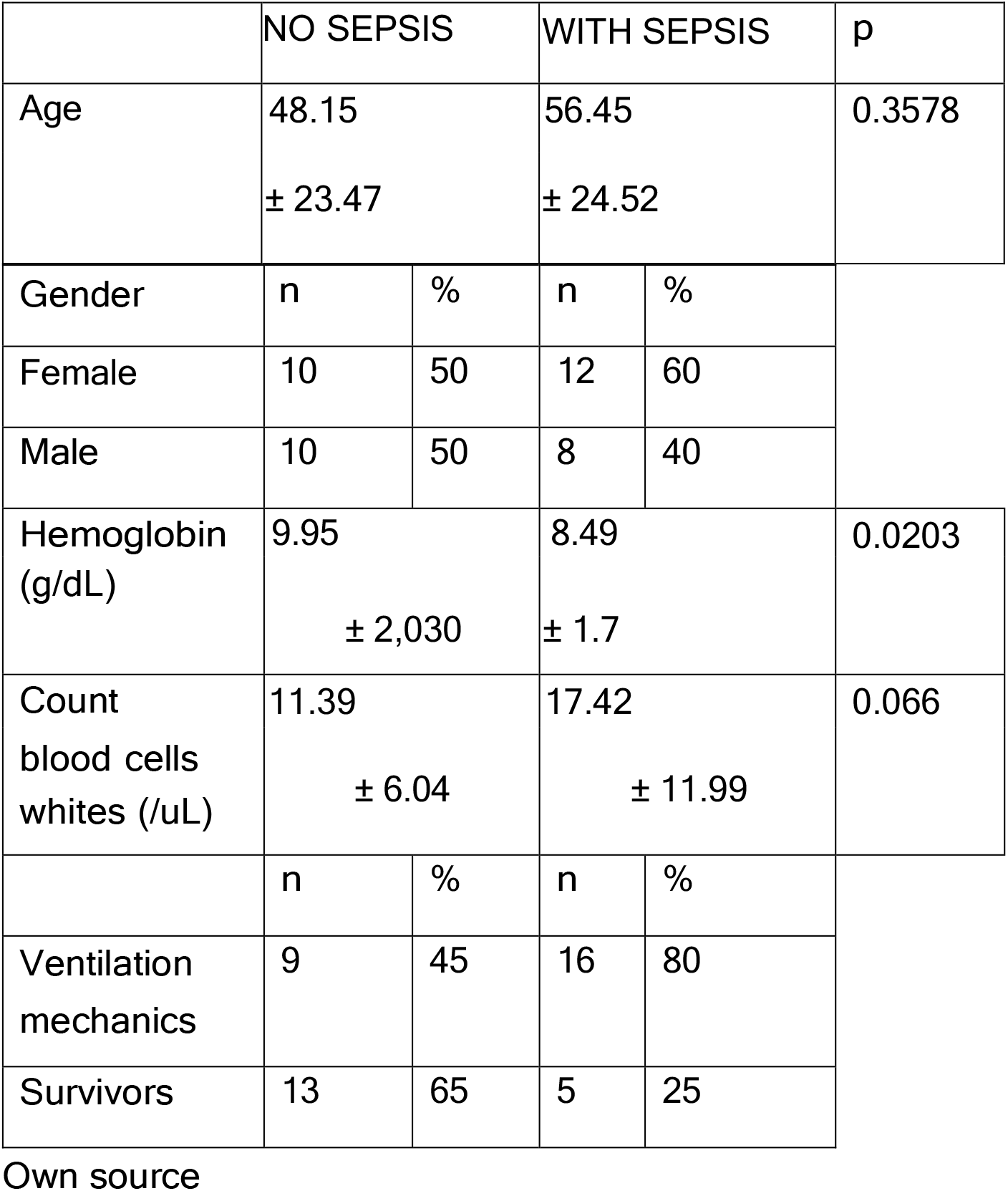
Distribution of participant groups by age, gender, and clinical characteristics.

|  | NO SEPSIS |  | WITH SEPSIS |  | p |
| --- | --- | --- | --- | --- | --- |
| Age | 48.15<br>± 23.47 |  | 56.45<br>± 24.52 |  | 0.3578 |
| Gender | n | % | n | % |  |
| Female | 10 | 50 | 12 | 60 |  |
| Male | 10 | 50 | 8 | 40 |  |
| Hemoglobin (g/dL) | 9.95<br>± 2,030 |  | 8.49<br>± 1.7 |  | 0.0203 |
| Count blood cells whites (/uL) | 11.39<br>± 6.04 |  | 17.42<br>± 11.99 |  | 0.066 |
|  | n | % | n | % |  |
| Ventilation mechanics | 9 | 45 | 16 | 80 |  |
| Survivors | 13 | 65 | 5 | 25 |  |

Own source

Regarding clinical and laboratory data, 80% of participants with sepsis were undergoing artificial mechanical ventilation, while only 45% of patients without sepsis were. Regarding deaths, 15 patients with sepsis (75%) died, while 13 in the group without sepsis survived (65%). The mean hemoglobin level in septic patients was 8.49 g/dL +/- 1.7 g, while in non-septic patients it was 9.95 g/dL (p=0.0203), with leukocyte counts of 17.42/uL +/- 11.99 g in septic patients and 11.39/uL in non-septic patients. See Table 1. Table 2 shows the most important clinical findings in cases of sepsis, the most frequent being leukocytes >12,000/uL (70%), heart rate >90 beats/min (65%), band count >10% (55%). The list of possible sources of sepsis is headed by the lungs (65%), followed by the abdomen (15%), kidneys (5%) and finally soft tissues (5%).

**Table 2.** Cytokine concentration and survival of patients with sepsis.

| **CYTOKINES vs SURVIVAL** |  |  |
| --- | --- | --- |
|  | **Survivors** | **No Survivors** |
| **IL-1** | 7.66 | 7.56 |
| | $\pm 0.73$ | $\pm 1.50$ |
| **IL-10** | 18.54 | 82.44 |
| | $\pm 2.50$ | $\pm 104.5$ |
| **TNF** | 30.63 | 27.71 |
| | $\pm 8.44$ | $\pm 9.79$ |

Source: own

The Shapiro-Wilk test showed that the data did not have a normal distribution; therefore, the Mann-Whitney U test was used to compare the median values of IL-1, IL-10, and TNF between septic and non-septic participants. The results showed no statistically significant difference between the medians of the cytokines IL-1 and TNF between the two groups (Figures 1A and 1B). However, there was a significant difference in IL-10 levels between the two groups, with IL-10 being higher in the sepsis group. Figure 1C. Regarding the levels of these cytokines according to the focus, a higher value of IL-10 was found in the pulmonary focus, as can be seen in Table 3.

**Table 3.**
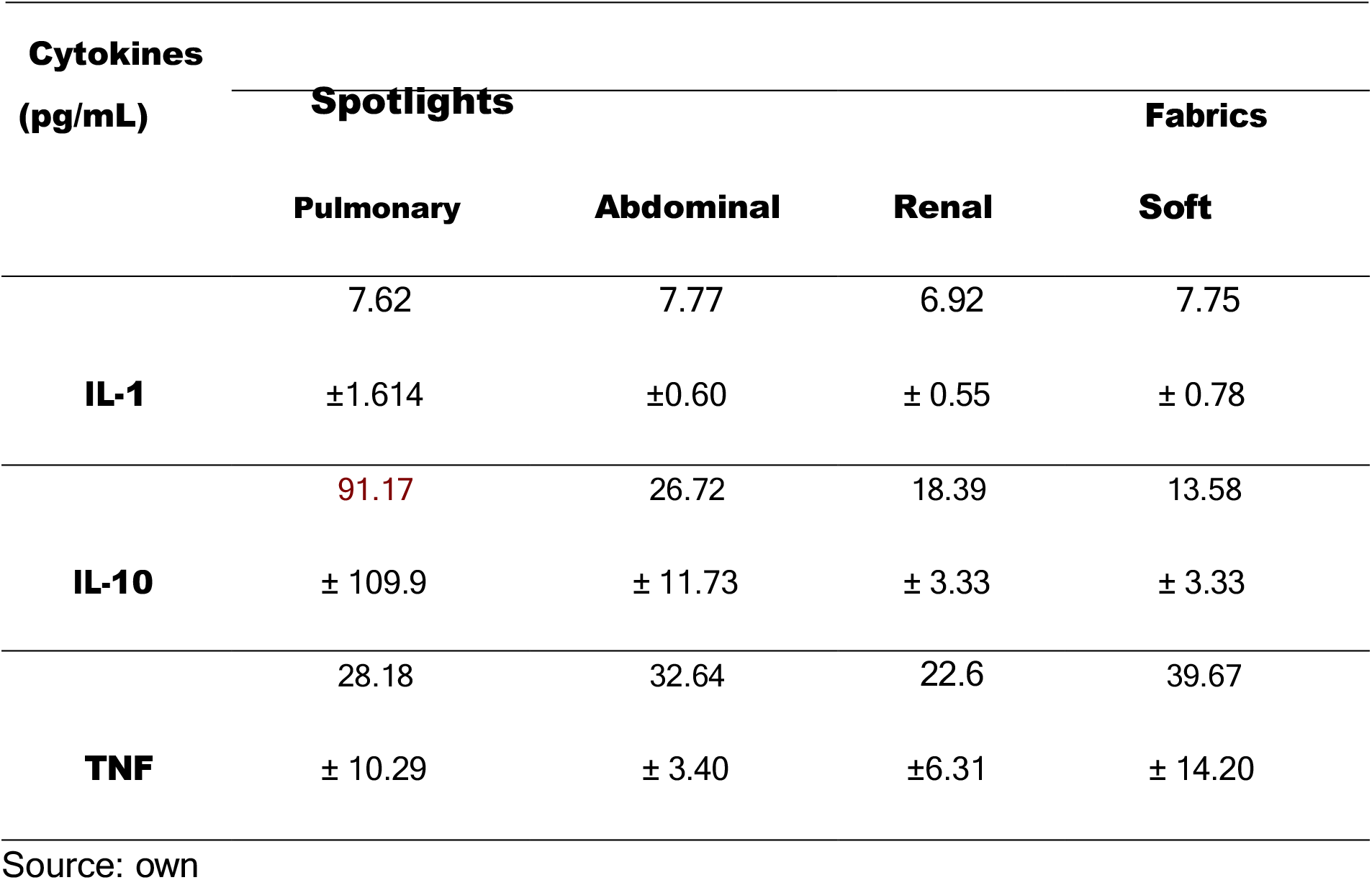
Plasma concentration of cytokines vs. sepsis-generating focus.

**Figure 1.**
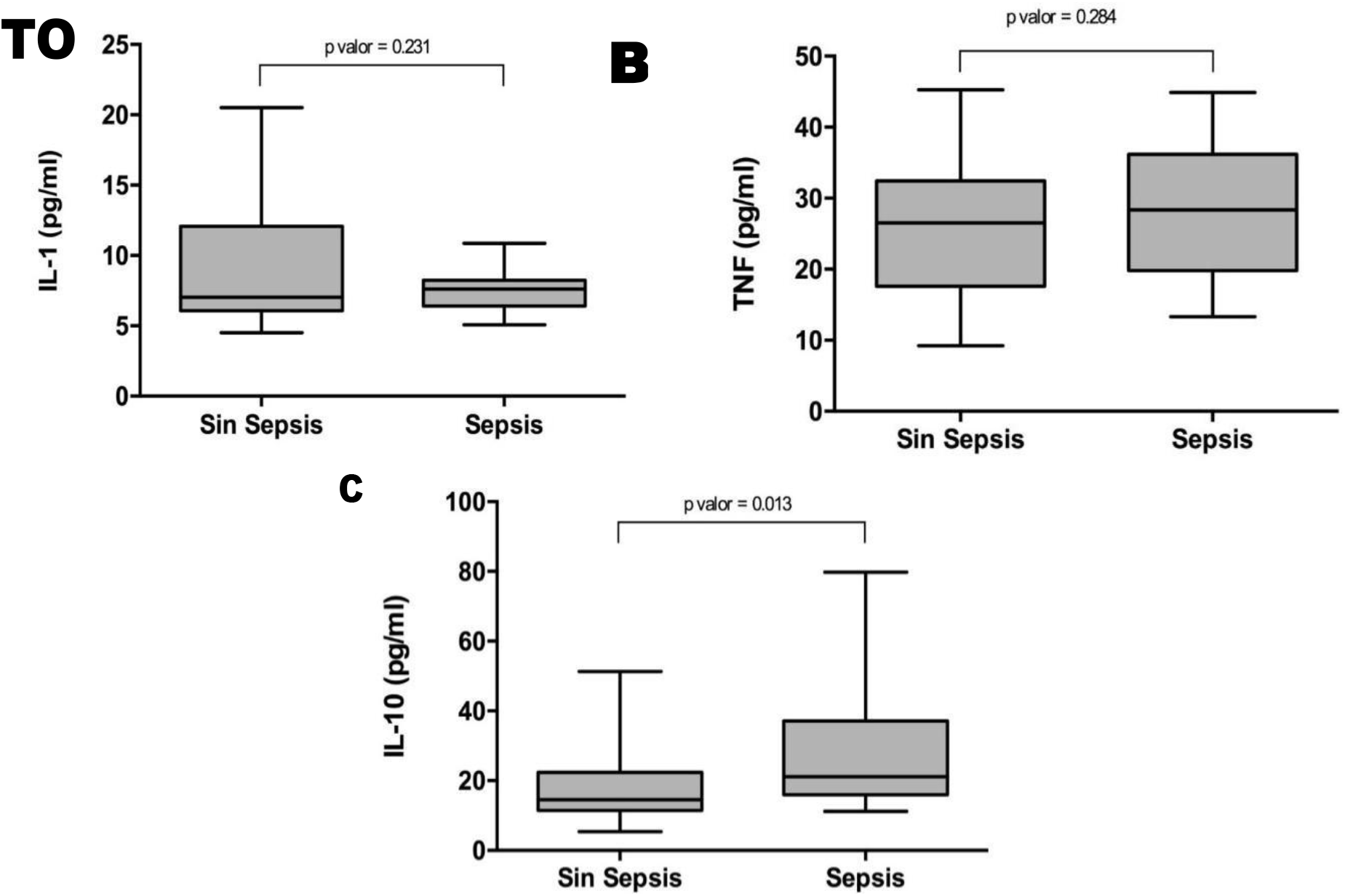
Comparison of cytokine levels between the groups of participants without sepsis vs. with sepsis. A. IL-1 levels, with no significant difference between the two groups, p=0.231. B. The graph shows TNF levels, with very similar values in both groups, p=0.264. C. IL-10 levels. The graph shows significantly higher IL-10 values in the sepsis group, p=0.013.

Regarding cytokine levels and mortality in participants, both IL-1 and TNF showed no significant correlation, with p values of 0.267 and p = 0.489, respectively. IL-10 did have a positive correlation (p = 0.001) with mortality.

## DISCUSSION

In this study, three cytokines that have commonly been classified as biomarkers of severity in sepsis were compared: IL-1, IL-10, and TNF (Faix, 2013; Reinhart, Bauer, Riedemann, & Hartog, 2012). It is important to note that in the group of patients diagnosed with sepsis, the majority received artificial mechanical ventilation, while in the group of patients without sepsis, less than half did, confirming that this invasive procedure increases the risk of a complication in the ICU, since in most of the patients in this study the main focus generating sepsis was the pulmonary focus, a fact directly related to the risk of death from sepsis.

The main findings of the study relate to a statistically significant difference between IL-10 levels in septic versus non-septic patients, being higher in the first group; while IL-1 and TNF were not.

showed significant differences. Similarly, considering the source of sepsis, participants with pulmonary sepsis showed higher IL-10 levels. Regarding mortality, IL-10 levels were statistically higher in non-survivors, while there were no differences in IL-1 and TNF levels.

Of special relevance among the findings is the positive relationship between IL-10 levels, disease severity and clinical outcome, taking into account that it is classified as an anti-inflammatory cytokine. The functions and importance of IL-10 and the importance of the balance between pro-inflammatory and anti-inflammatory cytokines in determining the degree and severity of the host’s immune response have been discussed extensively (14), with some authors even daring to assert the need to create drugs capable of increasing IL-10 levels and thus contribute to the suppression of inflammatory cytokines, as in the case of TNF, which is inhibited by IL-10 through a mechanism that stimulates the elongation of transcription by the cyclin-dependent kinase CDK 9.(15)(16).

The above is based on the premise that increased expression of proinflammatory cytokines plays a crucial role in the development of severe conditions such as sepsis, multiple organ failure, and even death. Therefore, it is essential that there be a balance in the kinetics of cytokine levels in the face of an infection. (16)

However, recently, contrary to what was believed, IL-10 has been linked to the severity of clinical symptoms, development of septic shock and a worse prognosis (17-19). For example, high levels of IL-10 have been found in non-survivors of septic shock. (18) Even in animal models, modulation of IL-10 expression has been shown to improve survival. This has led to a progressive rethinking of the Pathophysiological basis of sepsis. (20,21) concluding that IL-10 is involved in some type of immunosuppression that hinders the host’s ability to combat the pathogen that generates the primary infection, facilitating the acquisition of secondary lethal infections and even allowing the reactivation of latent viruses.

(22) (21). Similarly, in patients who died in the ICU due to sepsis, biochemical, flow cytometry and immunohistochemical findings compatible with immunosuppression have been found, compared to those who died from other causes (23). reprogramming (21). Another aspect to take into account is the existence of possible polymorphisms such as the SNP of the –1082A allele

There have been various attempts to explain why IL-10 levels increase, one of these is related to neutrophils, which are not usually responsible for the production of considerable amounts of IL-10, however, cases have been found in which due to their great capacity to adapt to a number of pathological and physiological conditions (24) such as septic processes, these can give rise to subgroups of neutrophils with immunosuppressive properties that lead to the production of significant amounts of IL-10 (21).

T cell changes in septic processes have also been reported, in which they adopt a tendency of hyporeactive proliferation towards a type 2 profile, with an increase in the production of IL-4 and IL-10. This increase in IL-10 leads to an increase in the proportion of regulatory T cells among the CD4+ T cell population, contributing to the state of latent immunosuppression, whose effect is related to a lower proliferation of effector T cells (25), suppression of innate immune cells, inhibition of monocytes and neutrophils, and induction of a phenomenon similar to endotoxin tolerance in NK cells. (19)

Another important finding in the present study is the fact that despite the existence of a hyperinflammatory process such as sepsis, no significant differences were found between the groups of participants with and without sepsis in pro-inflammatory cytokines such as IL-1 and TNF, these being one of the most extensively studied pro-inflammatory mediators in these cases and responsible for activating specific immune cells and consequently increasing the immune response.

In this regard, there are reports of patients with sepsis who have monocytes with a limited response capacity against endotoxin (lipopolysaccharide LPS), other TLR agonists and even other bacterial compounds (21), which some authors call endotoxin tolerance phenomenon. More specifically, these lymphocytes are characterized by decreased capacity to secrete pro-inflammatory cytokines such as TNF, IL-1, IL-6 and IL-12, while the secretion of anti-inflammatory mediators such as IL-1 receptor antagonist and IL-10 are normal and even improved (26).

Furthermore, reduced expression of human leukocyte antigen (HLA)-DR has been reported, resulting in a decrease in antigen presentation (19), possibly generated by changes in intracellular cell signaling that generate monocyte related to an increase in susceptibility to sepsis which also involves an increase in the production of IL-10 with a consequent immunosuppression (27).

## CONCLUSION

Given all the results found in the present study and those expressed by other authors, it is important that future studies include a greater number of variables that allow for a broader view of the immune response, such as the expression of HLA-DR, a greater number of cytokines, characterization of leukocyte populations, search for polymorphisms in genes related to IL-10, among others.

## Data Availability

All data produced in the present work are contained in the manuscript

